# Full Publication of Preprint Articles in Prevention Research: An Analysis of Publication Proportions and Results Consistency

**DOI:** 10.1101/2023.05.26.23290551

**Authors:** Isolde Sommer, Vincent Sunder-Plassmann, Piotr Ratajczak, Robert Emprechtinger, Andreea Dobrescu, Ursula Griebler, Gerald Gartlehner

## Abstract

**Introduction:** There is concern that preprint articles will lead to an increase in the amount of scientifically invalid work available. The objectives of this study were 1) to determine the proportion of prevention preprints published within 12 months, 2) to assess the consistency of the effect estimates and conclusions between preprint and published articles, and 3) to explore the reasons for the nonpublication of preprints.

**Methods:** We developed a web crawler to search the preprint server medRxiv for prevention studies posted from January 1 to September 30, 2020. An update search was conducted 12 months later. We dually screened the results for prevention articles and developed a scheme to classify changes in effect sizes and conclusions. We modeled the effect of a set of predictors on the proportion of preprints published in peer-reviewed journals. We also developed a questionnaire for authors of unpublished preprints.

**Results:** Of the 329 prevention preprints that met our eligibility criteria, almost half (48.9%) were published in a peer-reviewed journal within 12 months of being posted, with the median time being 5.3 months (range 0.1–11.3 months). While 27 out of 161 (16.8%) published preprints showed some change in the magnitude of the primary outcome effect estimate, 4.4% were classified as having a major change. The style or wording of the conclusion changed in 42.2%, while the content of the conclusion changed in 3.1%. Preprints on chemoprevention, with a cross-sectional design, and with public and noncommercial funding had the highest probabilities of publication. The main reasons for the nonpublication of preprints were journal rejection or lack of time.

**Conclusion:** The reliability of preprint articles for evidence-based decision-making is questionable. Less than half of the preprint articles on prevention research are published in a peer-reviewed journal within 12 months, and significant changes in effect sizes and/or conclusions are still possible during the peer-review process.

## Introduction

Preprints in health sciences have a relatively short tradition compared to other fields (e.g., physics, mathematics, biology) where researchers have been using preprint servers since the 1990s to distribute their research findings and ideas (1). With the founding of the medical preprint server medRxiv (www.medrxiv.org) in June 2019, preprints entered the field of medical and health research. The server’s popularity dramatically increased during the coronavirus disease 2019 (COVID-19) pandemic, which has boosted its use (2). These days, many journals (e.g., Elsevier [3], Springer [4], PLOS ONE [5], Lancet [6]) have introduced a preprint policy that allows or even encourages the sharing of preprints prior to peer-reviewed publication. Funding agencies such as Wellcome permit researchers to cite preprints in grant applications (7), and, in addition to encouraging preprint postings, they even require it if preprints being shared widely and rapidly results in a significant public health benefit (8).

Indeed, early and fast dissemination is the most appealing feature of preprints (9). Quick research sharing enables other researchers to build on early results, accelerating the research efforts necessary to overcome pressing health issues (10). There are concerns, however, that circumventing the peer-review process leads to an increase in the amount of scientifically invalid work (9). Some preprints have been cited widely in the press (10) and, without communicating the proper caution, there is a risk that their findings can be exaggerated by the media, while better-quality work could be ignored (11). According to a study in South Africa, 59% of news articles citing preprints failed to provide a statement of provisionality (12).

Various publications have discussed the validity of preprints, but despite the increasing amount of evidence on reporting and quality assurance, the available knowledge is still restricted to COVID-19, biomedical, or interdisciplinary research (13, 14, 15, 16, 17, 18, 19, 20, 21, 22, 23). No information exists on the use and validity of preprints in prevention research. Evidence from prevention research impacts community health and public health practice and informs public and policy decision-making every day, not only during emergent public health crises. It is therefore crucial to understand the validity of preprint results and conclusions in prevention research. The objectives of this study were

1. to determine the proportion of preprints that are published within 12 months of being added to medRxiv, overall and between different prevention types,
2. to assess the consistency of the effect estimates and conclusions between the preprint and published versions of prevention articles, and
3. to explore the reasons for the nonpublication of preprints in peer-reviewed journals.

## Materials and Methods

Our study protocol was registered in the Open Science Framework (OSF) (24). The overall project was a mixed-methods study and consisted of three parts: a text analysis, a qualitative interview study, and a survey study. Here, we report the results of the text analysis (i.e., objectives 1 and 2 of the larger overall project) as well as the results of the survey study, which we registered as an update to the original protocol in OSF (25).

### Text analysis

#### Data source and search strategy

We sampled studies from medRxiv (www.medrxiv.org). We developed a Python-based web crawler by analyzing the medRxiv website’s http responses using the Python packages requests (handling http) (26), BeautifulSoup (xml/html parsing) (27), and re (for extracting information from character strings related to prevention using regular expressions) (28) (see Table S1 for search string). We ran the web crawler on December 15, 2020 and let it search and extract information from prevention articles that had been posted on the medRxiv website (first run) from January 1, 2020 to September 30, 2020. It downloaded basic data about each identified preprint article: title, abstract, authors, version submission date, version history, download statistics, withdrawal information, funder, first author’s institutional affiliation, and information on the publication status (if published, new DOI, and the journal in which it appeared).

#### Inclusion and exclusion criteria for the study selection

We exported the information provided by the web crawler to Excel® for the abstract review. We dually screened all records identified by the web crawler against our eligibility criteria (see Table 1). Within the project, we used the working definition for prevention research established by the National Institute of Health Prevention Research (NIHR) Coordinating Committee (29). Using that definition, we included primary and secondary prevention studies that (1) identified and assessed risk and protective factors, (2) screened and identified individuals and groups at risk, (3) developed and evaluated interventions to reduce risk, (4) translated, implemented, and disseminated effective preventive interventions into practice, or (5) developed methods to support prevention research. We pilot-tested the abstract review with 50 records in the first web crawler round and amended the eligibility criteria where necessary. In case of uncertainty, we looked at the preprint’s full text and solved disagreements through discussion. We dually categorized each record according to the prevention categories (i.e., chemoprevention, counseling, immunization, screening, other primary prevention, other secondary prevention), whether COVID-19–related (i.e., yes or no), funding source (i.e., any funding vs. no funding, public or noncommercial funding, public and noncommercial funding, industrial funding, no funding), and study design (i.e., randomized controlled trial [RCT], cohort study, cross-sectional study, diagnostic study, ecological study, descriptive study, time series, before–after study, case control study, case series).

**Table 1:** Inclusion and exclusion criteria for the selection of preprints to be included in the text analysis

|  | **Inclusion criteria** | **Exclusion criteria** |
| --- | --- | --- |
| Topic | All prevention research (see the National Institute of Health Prevention Research [NIHR] Coordinating Committee's definition) (29), which can be categorized into: chemoprevention, counseling, immunization, screening, other primary prevention, other secondary prevention | Tertiary prevention, treatments, all other topics |
| Study designs | Clinical studies (including phase 2 | Modeling studies based on nonreal |
|  | trials), epidemiological studies, diagnostic studies | data, in vitro studies, qualitative studies, cost-effectiveness studies, all reviews (also systematic and rapid reviews), and basic research studies |
| Publication status | Preprints | All other publication types |
| Language | English | Other than English |
| Date posted on medRxiv | January 1, 2020 to September 30, 2020 | Later than September 30, 2020 |

#### Search for published preprint articles

To give every preprint a 12-month span to get published, we ran the web crawler again on October 5, 2021 and updated the information on publication status (second run). Because we did not want to rely entirely on the information provided by medRxiv regarding publication status, we manually searched Google® and GoogleScholar® for a published version of each unpublished preprint. If we still failed to identify a published version, we contacted the corresponding author of the unpublished preprint by email.

#### Data extraction and analysis

For preprints that were published in a peer-reviewed journal, we downloaded the article and performed further data extractions into a structured form using Excel®. One researcher extracted the following data, which was checked by a second researcher: the primary outcome effect estimate and conclusions regarding the primary outcome for both the preprint and peer-reviewed article, journal name, and publication date.

When we detected differences in the effect estimates or conclusions between the preprint and peer-reviewed article, two investigators independently classified these changes. We used the typology developed by Gartlehner et al. (30) to classify these changes but had to adapt it because of the range of effect estimates we identified. We considered the statistical significance of the primary outcome between the preprint and peer-reviewed article as having changed when at least one of the two effect estimates had a P-value that was deemed statistically significant in either the preprint or publication, and not statistically significant in the other. We assessed the magnitude of change in the effect estimate by applying the following categories to both dichotomous and continuous outcomes: no change, minor change (a relative change of up to 25 percentage points), and major change (a relative change of more than 25 percentage points).

For changes in the conclusion, we followed the categories suggested by Silagy et al. (31): no change, minor change (changes in style or wording that do not alter the substance or meaning of a section), and major change (changes that alter the substance or meaning of a section or alter the interpretation). The classifications of changes in the effect estimates and conclusions were done by one person and verified by a second person.

We retrieved the impact factor for each peer-reviewed journal from the Impact Factor List of 2019 provided by the Journal Citation Report (JCR) (32) and calculated the time until publication from the first appearance on medRxiv.

We used descriptive statistics and compared differences in publication characteristics and publication proportion between the different types of prevention articles. We used Bayesian methods to model the effect of a set of predictors on the proportion of peer-reviewed journal preprint publications as an outcome. The predictors included prevention type, whether COVID-19–related (yes/no), study design, and funding sources. We chose to use multiple individual models instead of one joint model for exploratory reasons, due to the lack of an underlying theory and to avoid issues associated with overadjustment and collider bias (33, 34, 35). The Bayesian modeling was conducted with Marcov Chain Monte Carlo methods via the brms package (36) and using restrictive priors. The intercept was suppressed. The statistical models were as follows:

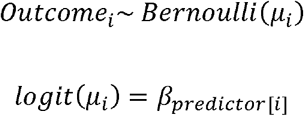

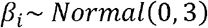

We conducted all analyses within the R environment (version 4.2.1). Additionally, we used the tidyverse (37) package, readxl (38), and tidybayes (39, 40) packages for data wrangling and creating the plots.

### Survey study

#### Survey development, participants, and procedure

We designed a questionnaire to explore the reasons for nonpublication in peer-reviewed journals and attitudes toward preprints in general. The questionnaire was developed in English and consisted of 11 items asking about the rationale behind deciding not publish the preprint or the reason(s) for and number of rejections as well as the estimated credibility of preprints, attitudes toward preprints, and demographic characteristics. The questionnaire was developed based on the results from the text analysis. The face vailidity was confirmed by the research team. Using the correspondence email address provided in the preprint, we sent the questionnaire to all corresponding authors of the prevention preprints identified in the text analysis that were not published at the start of the survey (n=152) between September 14 and November 14, 2022. We sent out several reminder emails to increase the response rate.

#### Ethics approval and compliance

The ethics committee of the University for Continuing Education, Krems, approved the survey study (EK GZ 28/2021-2024). Throughout the project, we adhered to the European Union data protection law (EU Regulation 2016/679). Participants were not reimbursed for their participation.

## Results

### Characteristics of the included preprints

In the first run, the web crawler identified 2238 preprints on medRxiv, of which we identified 594 as prevention research studies (26.5%). Among those, 329 were clinical, epidemiological, or diagnostic studies and met the inclusion criteria for our study selection (Fig. 1).

**Fig. 1:** Flow chart of the preprint selection process

Table 2 shows the characteristics of the included preprints. Of all the identified preprints, 73.6% (242/329) were on a topic related to COVID-19. Almost half focused on screening (46.2%), less than one-third (27.4%) were on other primary prevention topics, and about one-fifth were on immunization (20.1%). Few preprints fell into the other prevention categories (0.6% to 4.9%). The proportion of preprints receiving external funding was highest among the screening and chemoprevention preprints (59.9% and 62.5%). We identified most preprints as diagnostic (30.4%, 100/329) and cross-sectional studies (29.8%, 98/329) (Table S2). More than half had received external funding (48.6%, 160/329), mainly from public or noncommercial funding sources (29.8%, 98/329) (Table S3).

**Table 2:**
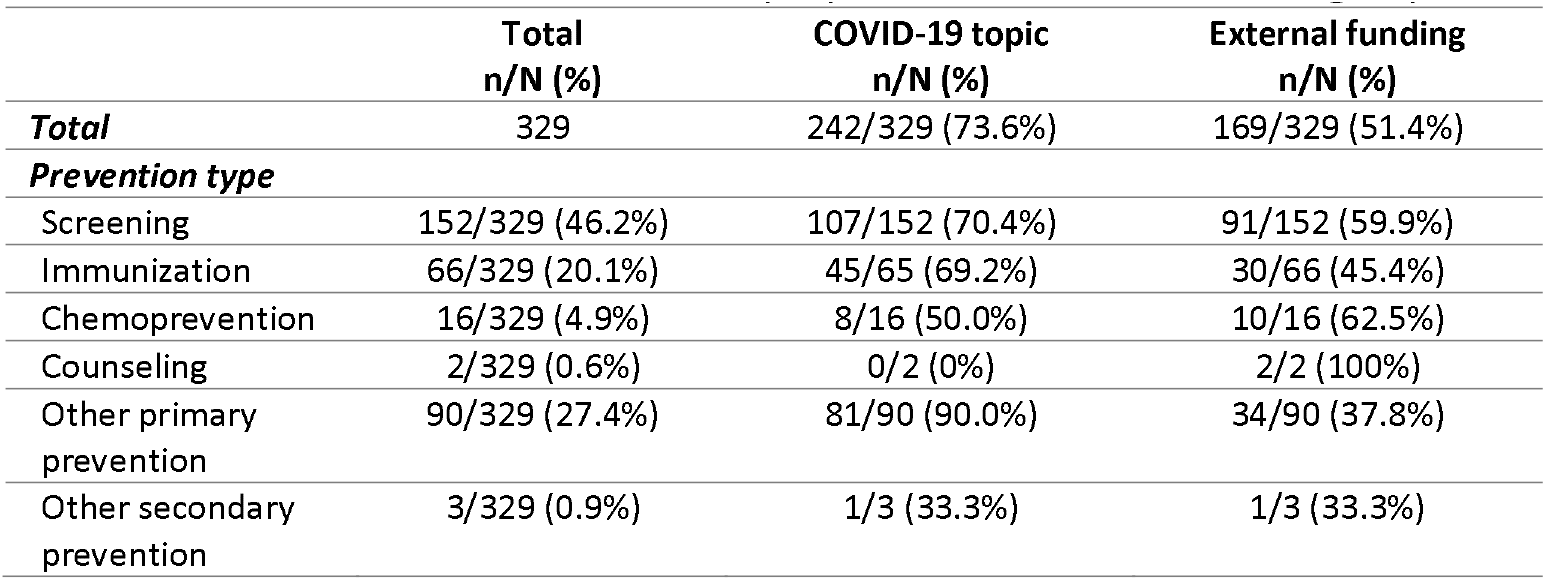
Characteristics of the included preprints in total and according to prevention type

### Proportion of published preprints

Of the 329 prevention preprints we identified, 161 (48.9%) were published in a peer-reviewed journal within 12 months of being uploaded to medRxiv (see Table 3). For published preprints, the median time from upload to journal publication was 5.3 months (range –0.1–11.3). COVID-19 studies were published more quickly than non-COVID studies (4.7 months; range -0.1^1^–11.3 vs. 7.0 months; range 1.3–11.3). The journals those preprints were published in had a median impact factor of 3.2 (range 0.2–74.7).

**Table 3:** Proportions of published preprints in total and according to prevention type

|  | Published within<br>12 months<br>n/N (%) | Median months until<br>publication (range) | Median impact factor of<br>journal where preprint<br>was published (range) |
| --- | --- | --- | --- |
| **Total** | 161/329 (48.9%) | 5.3 (-0.1*–11.3) | 3.2 (0.2–74.7) |
| COVID-19 | 116/242 (47.9%) | 4.7 (-0.1*–11.3) | 3.2 (0.2–74.7) |
| Non-COVID-19 | 45/87 (51.7%) | 7.0 (1.3–11.3) | 3.4 (0.67–59.1) |
| ***Prevention type*** |  |  |  |
| Screening | 58/152 (38.2%) | 5.9 (-0.1*–11.3) | 3.6 (0.2–53.4) |
| Immunization | 31/66 (47.0%) | 4.7 (1.0–10.8) | 3.6 (0.8–59.1) |
| Chemoprevention | 14/16 (87.5%) | 6.6 (0.3–10.6) | 3.2 (1.7–74.7) |
| Counseling | 2/2 (100%) | 4.7 (4.2–5.3) | 3.8 (2.1–5.6) |
| Other primary | 54/90 (60.0%) | 4.7 (0.2–10.8) | 2.9 (0.5–21.6) |
| prevention |  |  |  |
| Other secondary prevention | 2/3 (66.7%) | 5.1 (5.0–5.2) | 7.8 (5.4–10.2) |

\*Date of publication before first appearance on medRxiv

#### Proportions of published preprints according to prevention type, study design, and funding source

The proportions of published preprints differed between the prevention types, with preprints on screening having the lowest publication proportion (38.2%, 58/152) and those on chemoprevention having the highest proportion (87.5%, 14/16) (see Table 3). While the time from preprint to peer-reviewed journal publication showed little difference between prevention categories, the median impact factors of the journal preprints were highest in the other secondary prevention category, at 7.8 (range 5.4–10.2). For the study design, only about one-third of the diagnostic and ecological studies were published within 12 months (32.0%, 32/100 and 31.5%, 17/54), while 71.4% of the cross-sectional studies (70/98) were published within 12 months (see Table S4). The publication process took the least time for the cohort studies (median: 4.8; range 0.3–11.3) uploaded as preprints as compared to > 6 months for the diagnostic studies. According to the funding source, publicly and noncommercially funded studies had the highest proportions of published preprints (76.2%, 32/42) and impact factors (median 5.3, range 0.7–59.1). However, the publication of these studies also took the longest (median 6.2 months; range -0.1–9.8). In contrast, studies receiving no external funding were published faster (median 2.7 months, range 0.2–3.1) but in journals with lower impact factors (median 4.7, range 0.2– 11.0) (Table S5).

#### Publication predictors

Figures S1–S4 illustrate the effect of prevention type, COVID-19 (yes/no), study design, and funding sources as predictors of preprint publication. Based on our models, a higher probability of preprint publication was associated with chemoprevention topics (88%, 95% Credible Interval [Crl] 67.6 to 97.4), but only few studies were included in these categories. Cross-sectional study preprints (72%, 95% CrI 62.1 to 79.6) had the highest publication probability according to the study design. In terms of funding source, studies receiving public and noncommercial funding were the most likely to be published (77%, 95% CrI 62.3 to 87.3), while those with no external funding were the least likely (38%, 95% CrI 30.9 to 45.7). No difference was observed between COVID-19 and non-COVID-19 prevention studies (52%, 95% CrI 41.7 to 61.8 vs. 48%, 95% CrI 41.8 to 54.1).

### Consistency of the effect estimates

Out of 161 preprints that were published in a peer-reviewed journal, 16.8% (27/161) showed a change in the magnitude of the primary outcome effect estimate. This change was major (i.e., greater than 25%) in 4.4% studies (7/161) and minor (i.e., less than 25%) in 12.4% studies (20/161). Major changes comprised changes in the effect estimate magnitudes (1.9%, 3/161), in the primary outcomes (1.2%, 2/161), in the type of effect measure (0.6%, 1/161), and in assessment points (0.6%, 1/161). Among the 77 studies reporting statistical significance (47.8%), we did not observe any changes between the preprint and the peer-reviewed journal report, neither from statistically significant to nonsignificant nor vice versa.

### Consistency of the conclusions

The conclusions changed in 42.2% (68/161) of the articles after being published in a peer-reviewed journal compared to the preprint, mainly in terms of style or wording (39.1%, 63/161) (i.e., minor change). The content or meaning of the conclusion (i.e., major change) changed in 5 articles (3.1%).

### Survey of authors not publishing their preprint

We received a valid answer from 12 out of the 152 authors of preprints not published in a peer-reviewed journal within 12 months and with a valid email address (7.9% response rate). Eleven respondents were male, five were 50 years or older, and eight had more than 10 years of experience in research (Table S6). The reason most often given for nonpublication of the preprint in a peer-reviewed journal was rejection by at least one journal (58.3%, 7/12), followed by lack of time (25%, 3/12). Other reasons mentioned by one respondent each were that the preprint had received its attention, and that they had never intended to publish the preprint. Among those preprints that were submitted, 57.1% (4/7) got rejected 3–4 times. The official reasons given by journals for rejecting the preprints were manyfold and included lack of novelty (n=3), too few figures/tables (n=2), and not meeting the journal’s scope (n=2), among others. Two-thirds of the preprints did not receive external funding (66.7%, 8/12). The reasons most often indicated for uploading the preprint were sharing the results with the community (n=11), immediate/fast publication (n=6), and increased visibility of work (n=6). All the results are presented in Table S7.

## Discussion

### Main findings

To the best of our knowledge, this study is the first to provide a thorough analysis of the publication proportion and consistency of the effect estimates and conclusions of preprints and their subsequent publications in prevention research. Almost half of the prevention preprints (48.9%) were published in a peer-reviewed journal after 12 months of being uploaded to medRxiv, with the median time from upload to publication being 5.3 months (range –0.1*–11.3 months). About half of the prevention preprints were on screening (46.2%), a quarter on other primary prevention topics (27.4%), and one-fifth on immunization (20.1%). The results from the modeling analysis indicate that preprints on chemoprevention and cross-sectional studies or those with public and noncommercial funding had the highest probability of publication within their categories. Preprint authors who did not publish their results in peer-reviewed journals mentioned journal rejections followed by lack of time as the main reasons for nonpublication.

We detected a change in the magnitude of the effect estimates in 27 out of the 161 preprints that were published in a peer-reviewed journal (16.8%), but most were minor changes. Although changes in the magnitude of the effect estimate were predominately minor and did not appear very often, they still warrant caution for the use of preprints in decision-making in the prevention field. If 7 out of 161 articles had a major change in the magnitude of the effect estimate, every 23^rd^ article is affected. In addition, it must be considered that as yet we have no knowledge of the quality of unpublished results. We found changes in the conclusions in 42.2% of the preprints that were published within 12 months, but mostly in terms of style or wording, and only in 5 out of 161 articles was the content of the conclusion changed (3.1%). It therefore seems very sensible that medRxiv has issued a warning on the main page of their website that preprints should not be relied on to guide clinical practice or health-related behavior and should not be reported as established information by news media (41). A definite assessment of the credibility of preprints will be possible when the reasons for not publishing them are fully understood.

### Relation of the study’s findings to previous work

Several studies have centered their work on the publication proportions of preprint articles. Recently, studies focusing on COVID-19 preprints reported proportions of 5.7% to 55.3% preprints published within the study periods (5 to 18 months), with a median time to publication between 2.3 and 5.9 months (13, 18, 19, 20). A third of the preprint articles uploaded to bioRxiv, a preprint server for biology research (https://www.biorxiv.org/), before 2017 did not get published as peer-reviewed articles (21).

The proportion of preprints published in our study (48.9%) is close to that reported in the studies by Otridge et al. (18) and Zeraatkar et al. (19), but these are not directly comparable given the different study periods (12 months vs. 16 and 18 months). Although we did not find that whether an article was COVID-19–related predicted the publication proportion, our findings demonstrate that such articles are published more quickly. An analysis of COVID-19 articles from January to June 2020 showed that peer review was accelerated for COVID-19 articles but decelerated for non-COVID-19 articles because all resources were pushed toward COVID-19 (42). Other studies found more COVID-19 studies published within their study period than non-COVID-19 studies (22, 43).

Like in our study, Zeraatkar et al. (19) investigated predictors of preprint publication but came up with different results. They found that preprints were more likely to be published if they received government funding. Our study identified public and noncommercial funding as the strongest predictor for publishing but used different categories for funding source. We further found that chemoprevention and cross-sectional studies had the highest publication probabilities within their categories. To fully understand which factors predict preprint publication, it is important to undertake a larger, more detailed analysis of preprints.

As for the changes in effect estimates and conclusions, our study’s findings largely mirror those of other studies reporting on the consistency between preprints and subsequently published articles (14, 15, 16, 19). For example, Bero and colleagues (14) did not find large discrepancies in results reporting or the presence of spin between COVID-19 interventional and observational preprints and publications, but small changes were frequent. Another study using a stricter classification scheme (important change in any effect estimate by ≥10% and/or change in significance level) classified 21% of COVID-19 intervention trials as having an important change from preprint to peer-reviewed article (44), which is much higher than in our study, as we found that only 4.4% of studies had a major change in the effect estimates.

The debate around the credibility of preprints opens the discussion on the quality of peer review. In a study on articles published in bioRxiv and in PubMed-indexed journals, peer-reviewed articles had a higher reporting quality than preprints, particularly in how clearly the titles and abstracts presented the main findings and how easy it was to locate relevant reporting information (45). Changes in reporting from preprints to peer-reviewed versions did not correlate with the journal’s impact factor, a finding that resonates with the range of impact factors in our study (median 3.2, range 0.2–74.7). Garcia-Costa et al. (46), who assessed the developmental function of peer review in their study, concluded that taking more time to deliver a peer review increases a manuscript’s rigor and value through constructive feedback. Given the increasing challenges journal editors face in recruiting qualified peer-reviewers, the concept of community-based peer review seems appealing. Community-based peer review of posted preprints allows quick scrutiny of research and fast withdrawals when concerns are raised (47). However, it is unclear how widely community-peer review is used and what its impact on the quality of preprints is.

### Strengths and limitations of the study

The strengths of this study include the use of a web crawler, which allowed us to automatically screen the medRxiv server, identify preprints in prevention research, and extract relevant information. The web crawler also made it easy to track the publication status of these preprints after 12 months. Another strength is the dual screening, dual data extraction, and dual categorization of the prevention preprints. Finally, we performed a thorough assessment of the included preprint prevention articles, ranging from publication proportion to overall changes in effect size and conclusion and the identification of publication predictors. We are very confident not to have missed any preprint publications within the study period, as we contacted all preprint authors for assurance. Another strength is the additional survey among the preprint authors whose preprints were not published after 12 months to gain further insight on the reasons for nonpublication. A limitation, however, is the low response rate (7.9%), which precluded us from making generalized conclusions.

The limitations of the study include the small sample size to analyze differences across the prevention categories, funding sources, and study designs. We further made decisions on the categorization of prevention research, study design, and funding source based only on the information provided in the abstracts; therefore, it is possible that we miscategorized some of them. While the correct classification was important for identifying the predictors, it did not have an influence on the assessment of the changes in the effect estimates and conclusions between the preprint and peer-reviewed article.

## Conclusions

This study expands our knowledge that preprints on prevention research topics have few major changes in the effect estimates and conclusions after undergoing the peer-review process. Although, at first sight, these changes appear in a small number of preprints, still, every 23^rd^ article is affected. We therefore warrant caution in using preprints of prevention research in decision-making.

## Data Availability

All relevant are available from the OSF database (https://osf.io/cnkdw)

## Acknowledgments

We would like to thank Florian Nehonsky for developing the web crawler, Irma Klerings for providing the search string, Andrea Trampert for help with the literature screening, Emma Persad for second-checking the effect size and conclusion classifications, and Manuela Müllner for administrative support.

## Author contributions

Conceptualization: IS, GG, UG, AD

Investigation: IS, VSP, PR, AD, UG

Formal analysis: RE

Writing – Original Draft Preparation: IS

Writing – Review & Editing: IS, GG, UG, VSP, PR, RE, AD

## Supporting information

Table S1: Search string for the web crawler

Table S2: Characteristics of the included preprints according to study design

Table S3: Characteristics of the included preprints according to funding source

Table S4: Proportions of published preprints according to study design

Table S5: Proportions of published preprints according to funding source

Figure S1: Prevention type as a predictor of preprint publication (probability expressed as %, 95% credible intervals)

Figure S2: COVID-19 (yes/no) as a predictor of preprint publication (probability expressed as %, 95% credible intervals)

Figure S3: Study design as a predictor for preprint publication (probability expressed as %, 95% credible intervals)

Figure S4: Funding source as a predictor for preprint publication (probability expressed as %, 95% credible intervals)

Table S6: Sample characteristics of authors of nonpublished preprints (n=12)

Table S7: Survey results (n=12)

