## Supplementary material for "Full Publication of Preprint Articles in Prevention Research: An Analysis of Publication Proportions and Results Consistency"

**Appendix**

Table S1: Search string for webcrawler

*Medarxiv:* December 15, 2020 and October 5, 2021

prevention OR preventive OR immunisation* OR immunization* OR vaccination* OR counselling OR counseling OR "health promotion" OR "health education" OR "behaviour change" OR "behavior change" OR "lifestyle intervention" OR screening OR "early detection" OR "early diagnosis" OR chemoprevention OR chemoprophylaxis OR prophylaxis OR prophylactic OR "risk reduction" OR "reduce the risk" OR "reducing the risk"

Table S2: Characteristics of included preprints according to study design

|  | Total  n/N (%) | Covid-19 topic  n/N (%) | External funding  n/N (%) |
| --- | --- | --- | --- |
| *Study design* | | | |
| Diagnostic study | 100/329 (30.4%) | 73/100 (73.0%) | 60/100 (60.0%) |
| Cross-sectional study | 98/329 (29.8%) | 72/98 (73.5%) | 45/98 (45.9%) |
| Ecological study | 54/329 (16.4%) | 46/54 (85.2%) | 16/54 (29.6%) |
| Descriptive study | 25/329 (7.6%) | 22/25 (88.0%) | 9/25 (36.0%) |
| Cohort study | 25/329 (7.6%) | 12/25 (48.0%) | 15/25 (60.0%) |
| Before-after study | 13/329 (4.0%) | 11/13 (84.6%) | 3/13 (23.1%) |
| RCT | 7/329 (2.1%) | 4/7 (57.1%) | 7/7 (100%) |
| Time series | 5/329 (1.5%) | 2/5 (40.0%) | 3/5 (60.0%) |
| Case control study | 1/329 (0.3%) | 0/1 (0%) | 1/1 (100%) |
| Case series | 1/329 (0.3%) | 0/1 (0%) | 1/1 (100%) |

RCT, randomized controlled trial;

Table S3: Characteristics of included preprints according to funding source

|  | Total  n/N (%) | Covid-19 topic  n/N (%) |
| --- | --- | --- |
| *Study design* | | |
| Public or non-  commercial | 98/329 (29.8%) | 63/98 (64.3%%) |
| Public and non-  commercial | 42/329 (12.2%) | 17/42 (40.5%) |
| Industry | 20/329 (6.1%) | 11/20 (55.0%) |
| No external funding | 160/329 (48.6%) | 144/160 (90.0%) |
| Not reported | 9/329 (2.7%) | 7/9 (77.8%) |

Table S4: Proportions of published preprints according to study design

|  | Published within  12 months  n/N (%) | Median impact factor (range) | Median months until  publication (range) |
| --- | --- | --- | --- |
| *Study design* | | | |
| Diagnostic study | 32/100 (32.0%) | 5.7 (0.2 – 53.4) | 6.2 (1.4 – 11.1) |
| Cross-sectional study | 70/98 (71.4%) | 3.2 (0.5 – 25.1) | 5.1 (-0.1* – 10.8) |
| Ecological study | 17/54 (31.5%) | 5.0 (0.8 – 35.5) | 5.2 (1.0 – 10.8) |
| Descriptive study | 13/25 (48%) | 2.9 (0.2 – 39.9) | 5.5 (2.5 – 10.6) |
| Cohort study | 13/25 (52.0%) | 3.2 (1.4 – 22.7) | 4.8 (0.3 – 11.3) |
| Before-after study | 4/13 (30.8%) | 2.6 (2.2 – 6.8) | 3.5 (0.2 – 7.0) |
| RCT | 7/7 (100%) | 8.3 (2.4 – 74.7) | 4.6 (0.9 – 10.8) |
| Time series | 3/5 (60.0%) | 2.7 (2.0 – 4.3) | 10.4 (0.9 – 10.9) |
| Case control study | 1/1 (100%) | 3.1 (NA) | 5.2 (NA) |
| Case series | 1/1 (100%) | 11.1 (NA) | 7.3 (NA) |

*Date of publication before first appearance on medRxiv

RCT, randomized controlled trial; NA, not applicable;

Table S5: Proportions of published preprints according to funding source

|  | Published within  12 months  n/N (%) | Median impact factor (range) | Median months until  publication (range) |
| --- | --- | --- | --- |
| *Funding source* | | | |
| Public or non-  commercial | 54/98 (55.1%) | 3.2 (0.9 – 53.4) | 5.3 (0.5 – 11.2) |
| Public and non-  commercial | 32/42 (76.2%) | 5.3 (0.7 – 59.1) | 6.2 (-0.1 – 9.8) |
| Industry | 10/20 (50.0%) | 4.3 (2.4 – 74.7) | 5.7 (0.9 – 11.0) |
| No external funding | 61/160 (38.1%) | 2.7 (0.2 – 13.1) | 4.7 (0.2 – 11.0) |
| Not reported | 4/5 (80.0%) | 2.2 (1.1 – 2.3) | 5.1 (1.6 – 6.3) |

Figure S1: Prevention type as predictor for publication of preprints (probability expressed as %, 95% credible intervals)

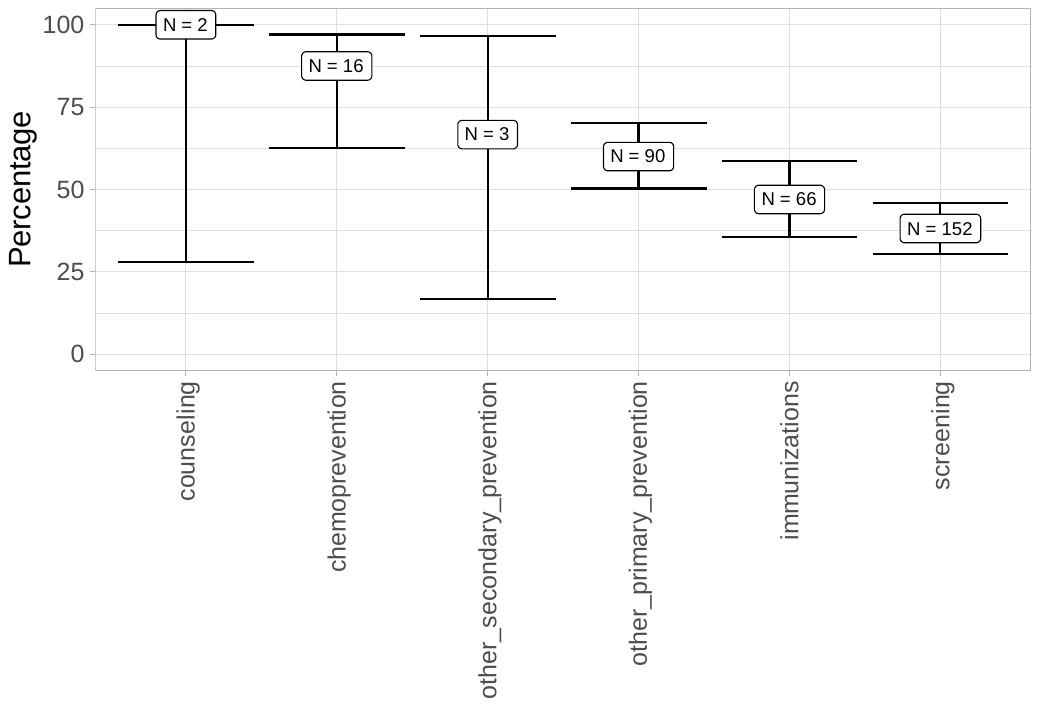

Figure S2: COVID-19 yes/no as predictor for publication of preprints (probability expressed as %, 95% credible intervals)

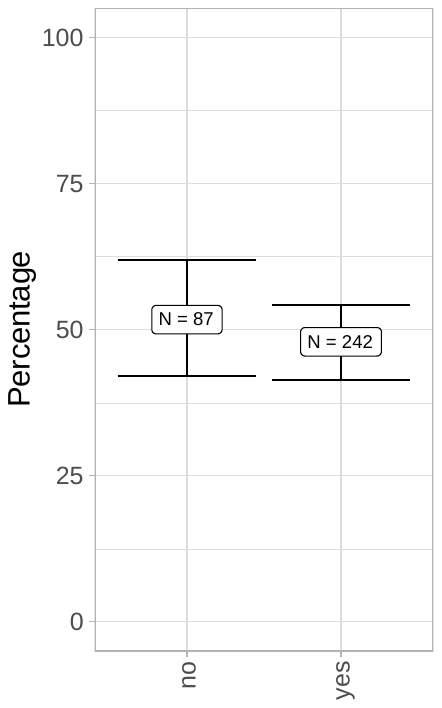

Figure S3: Study design as predictor for publication of preprints (probability expressed as %, 95% credible intervals)

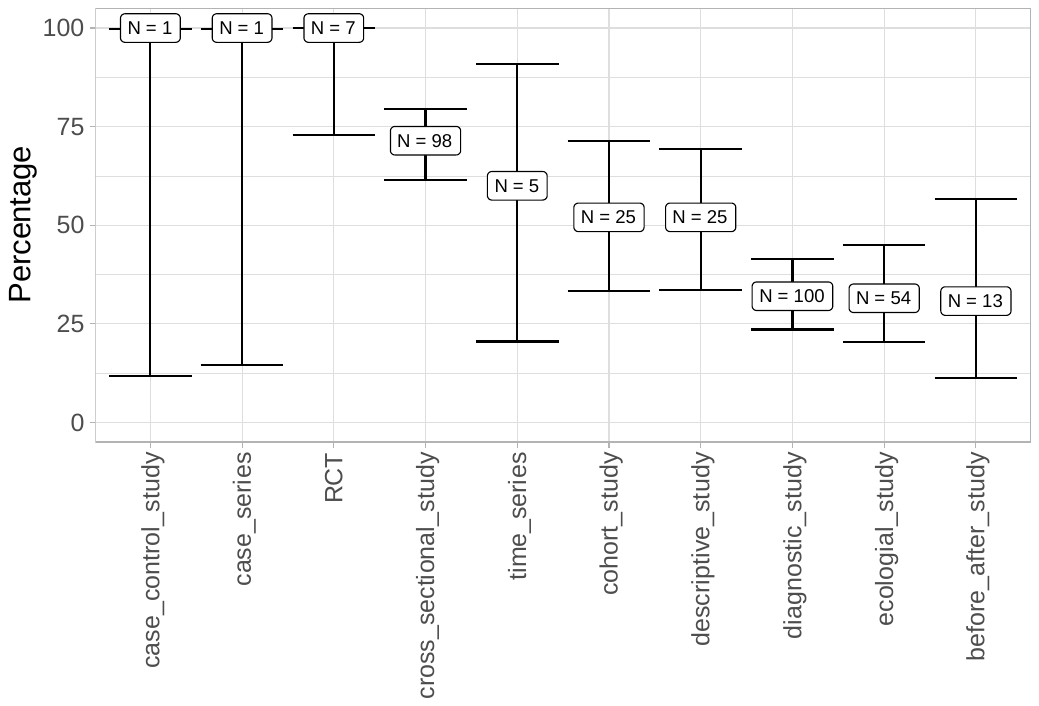

Figure S4: Funding source as predictor for publication of preprints (probability expressed as %, 95% credible intervals)

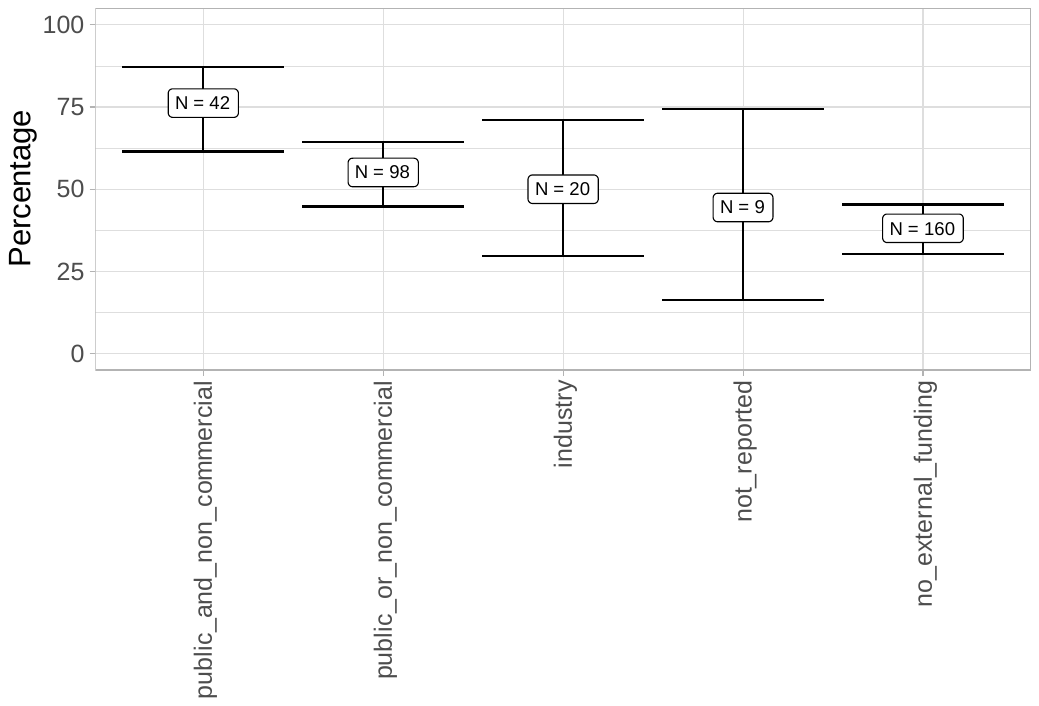

Table S6: Sample characteristics of authors of non-published preprints (n=12)

| **Author characteristics** | Percentage | Numbers |
| --- | --- | --- |
| **Age in years** | | |
| 20-29 | 8.3% | 1 |
| 30-39 | 33.3% | 4 |
| 40-49 | 16.7% | 2 |
| 50-59 | 25.0% | 3 |
| 60-69 | 8.3% | 1 |
| 70-79 | 8.3% | 1 |
| **Gender** | | |
| Male | 91.7% | 11 |
| Female | 8.3% | 1 |
| **Country of work** | | |
| China | 8.3% | 1 |
| India | 25.0% | 3 |
| Nigeria | 8.3% | 1 |
| Pakistan | 8.3% | 1 |
| Philippines | 8.3% | 1 |
| UK | 8.3% | 1 |
| USA | 33.3% | 4 |
| **Current position** | | |
| Student | 8.3% | 1 |
| Assistant/Associate Professor | 16.7% | 2 |
| Professor | 16.7% | 2 |
| Researcher in academia/industry/  non- academic organizations | 33.3% | 4 |
| Clinician | 8.3% | 1 |
| Other:   1. Chief Medical Officer of Health of a district 2. Limited Liability Company member | 16.7% | 2 |
| **Years of experience in research** | | |
| 1-10 | 33.3% | 4 |
| 11-20 | 41.7% | 5 |
| 21-30 | 16.7% | 2 |
| >30 | 8.3% | 1 |

Table S7: Survey results (n=12)

| Question item | Percentage | Numbers |
| --- | --- | --- |
| **Reasons for not publishing the preprint in a peer-reviewed journal within one year after being uploaded to *medRxiv*** | | |
| Lack of time | 25.0 | 3 |
| Rejection by ≥ 1 journal | 58.3 | 7 |
| Other:   1. It has received its due attention 2. No intention of peer-reviewed publication. | 16.7 | 2 |
| **Number of times the paper was rejected (if it was rejected) (n=7)** | | |
| 1-2 | 28.6 | 2 |
| 3-4 | 57.1 | 4 |
| 5-6 | 14.8 | 1 |
| **Official reason for rejection of the paper in a peer-reviewed journal (multiple answers possible)** | | |
| Too few figures/tables | NA | 2 |
| Journal’s scope was not met | NA | 2 |
| Lack of novelty | NA | 3 |
| Inappropriate study design | NA | 1 |
| Writing style | NA | 1 |
| Poor language quality | NA | 1 |
| Too many incoming manuscripts | NA | 2 |
| Other:   1. Word count exceeded 2. No reason given 3. Not sufficient interest | NA | 4  1  2  1 |
| **Funding source of non-published preprint** | | |
| Commercial (industrial) | 8.3 | 1 |
| Governmental | 25.0 | 3 |
| Non-governmental | 0 | 0 |
| Mixed | 0 | 0 |
| No external funding | 66.7 | 8 |
| **Reasons for uploading preprint (multiple answers possible)** | | |
| Sharing results with the community | NA | 11 |
| Immediate/fast publication | NA | 6 |
| Increased visibility of work | NA | 6 |
| Other:   1. COVID | NA | 1 |
| **Numbers of preprints having uploaded in the past** | | |
| 1 | 25.0 | 3 |
| 2-5 | 50.0 | 6 |
| More than 5 | 25.0 | 3 |
| **Number of preprints having published in a journal in the past** | | |
| 0 | 33.3 | 4 |
| 1 | 33.3 | 4 |
| 2-5 | 16.7 | 2 |
| More than 5 | 16.7 | 2 |
| **“Preprints have an appropriate level of credibility.”** | | |
| Strongly agree | 16.7 | 2 |
| Agree | 33.3 | 4 |
| Undecided | 33.3 | 4 |
| Disagree | 16.7 | 2 |
| Strongly disagree | 0.0 | 0 |
| **Do you cite preprints of other authors who do not get published in peer-reviewed journals in your manuscripts?** | | |
| Often | 25.0 | 3 |
| Sometimes | 41.7 | 5 |
| Seldom | 16.7 | 2 |
| Never | 16.7 | 2 |

NA, not applicable
